# The impact of burn trauma on glycocalyx derangement

**DOI:** 10.1101/2024.09.02.24312926

**Authors:** Hannes Kühtreiber, Daniel Bormann, Melanie Salek, Thomas Haider, Caterina Selina Mildner, Marie-Therese Lingitz, Clemens Aigner, Christine Radtke, Hendrik Jan Ankersmit, Michael Mildner

## Abstract

Burn injuries often lead to severe complications, including acute respiratory distress syndrome (ARDS), driven in part by systemic inflammation and glycocalyx disruption. In this study, we analyzed the sera of 28 patients after burn trauma and utilized transcriptomic analyses to decipher the impact of burn injury on glycocalyx derangement. We observed significant upregulation of immune cell-derived degrading enzymes, particularly matrix metalloproteinase-8 (MMP8), which correlated with increased immune cell infiltration and glycocalyx derangement. Serum analysis of burn patients revealed significantly elevated levels of shed glycocalyx components and MMP8, both correlating with the presence of inhalation injury. Consequently, treatment of human in vitro lung tissue models with MMP8 induced significant glycocalyx shedding in both, the endothelium and epithelium. Together our data suggest MMP8 as a contributor of glycocalyx disruption and lung injury post-burn.

## Brief communications text

Burn injuries predominantly affect the skin and lungs and are associated with high morbidity and mortality ^1^ The standard management of burn injuries includes fluid resuscitation, wound care, and surgical interventions such as debridement or skin grafting. These treatments are complemented by pain management and continuous rehabilitation to support both physical and psychological recovery ^2^. Despite advancements in care, severe complications like multiple organ failure, sepsis, and respiratory failure continue to significantly impact patient outcomes, contributing to high mortality rates ^1^. Our previous studies have highlighted the role of systemic inflammation and neutrophils in the complications following burn injuries ^3, 4^. While neutrophils are crucial for the innate immune response and tissue debridement, their overactivation post-burn results in substantial collateral tissue damage. These activated neutrophils are able to migrate to the lungs, thereby contributing to tissue damage through the induction of NETosis, degranulation, and the production of reactive oxygen species (ROS) ^5^. These processes can exacerbate complications like acute respiratory distress syndrome (ARDS) ^6^.

Given this context, the glycocalyx, a dynamic pericellular matrix composed primarily of proteoglycans and glycosaminoglycans (GAGs), plays a critical role in maintaining vascular and lung tissue integrity ^7^.This matrix undergoes continuous remodeling, with its degradation often driven by factors such as mechanical stress, matrix metalloproteinases (MMPs) ^8^, and ROS in response to pathological conditions and inflammation ^9^. Disruption of the glycocalyx, particularly in the lung, is linked to severe outcomes such as ARDS, as evidenced by elevated serum levels of syndecan-1 (SDC1) in patients with septic shock and non-pulmonary sepsis ^10^. Despite the substantial focus on the endothelial glycocalyx in the lungs, the glycocalyx of the alveolar epithelium remains underexplored. Direct lung injury can lead to the shedding of the alveolar epithelial glycocalyx, which in turn impairs surfactant function, correlating with the severity and duration of respiratory complications. This impairment is associated with atelectasis and reduced lung compliance ^11^.

In this study, we aimed to explore the impact of burn injury on glycocalyx disruption, with a particular focus on the role of degrading enzymes derived from innate immune cells. Our goal was to provide new insights into the processes underlying glycocalyx derangement and subsequent lung damage following burn injury.

To assess the impact of burn trauma on glycocalyx components and degrading enzymes, we re-analyzed publicly available single-cell RNA sequencing (scRNA-seq) data from a murine burn injury model ^12^. Unbiased clustering analysis identified 12 distinct cell populations, which were further annotated using established marker genes (Fig. 1a, Fig. S1a). Our analysis revealed a dynamic interplay between glycocalyx components and degrading enzymes following burn injury. We observed a notable decrease in the overall expression of glycocalyx-related mRNAs by day 3 post-injury, followed by a gradual increase, suggesting a dynamic reparative process (Fig. 1b). Simultaneously, the expression of degrading enzymes peaked on day 3, particularly within innate immune cell populations (Fig. 1c). This peak coincided with a significant rise in the relative proportion of innate immune cells (Fig. S1b). Notably, the glycocalyx module score increased primarily in the fibroblast population, while expression in endothelial cells remained largely unchanged. These findings suggest that endothelial cells, despite being common targets of glycocalyx damage post-burn, exhibit a less pronounced repair response compared to other cell types. This endothelial damage is particularly concerning in pulmonary complications, where it exacerbates lung injury by compromising vascular integrity and exposing adhesion molecules that facilitate the diapedesis of immune cells.

**Figure 1.**
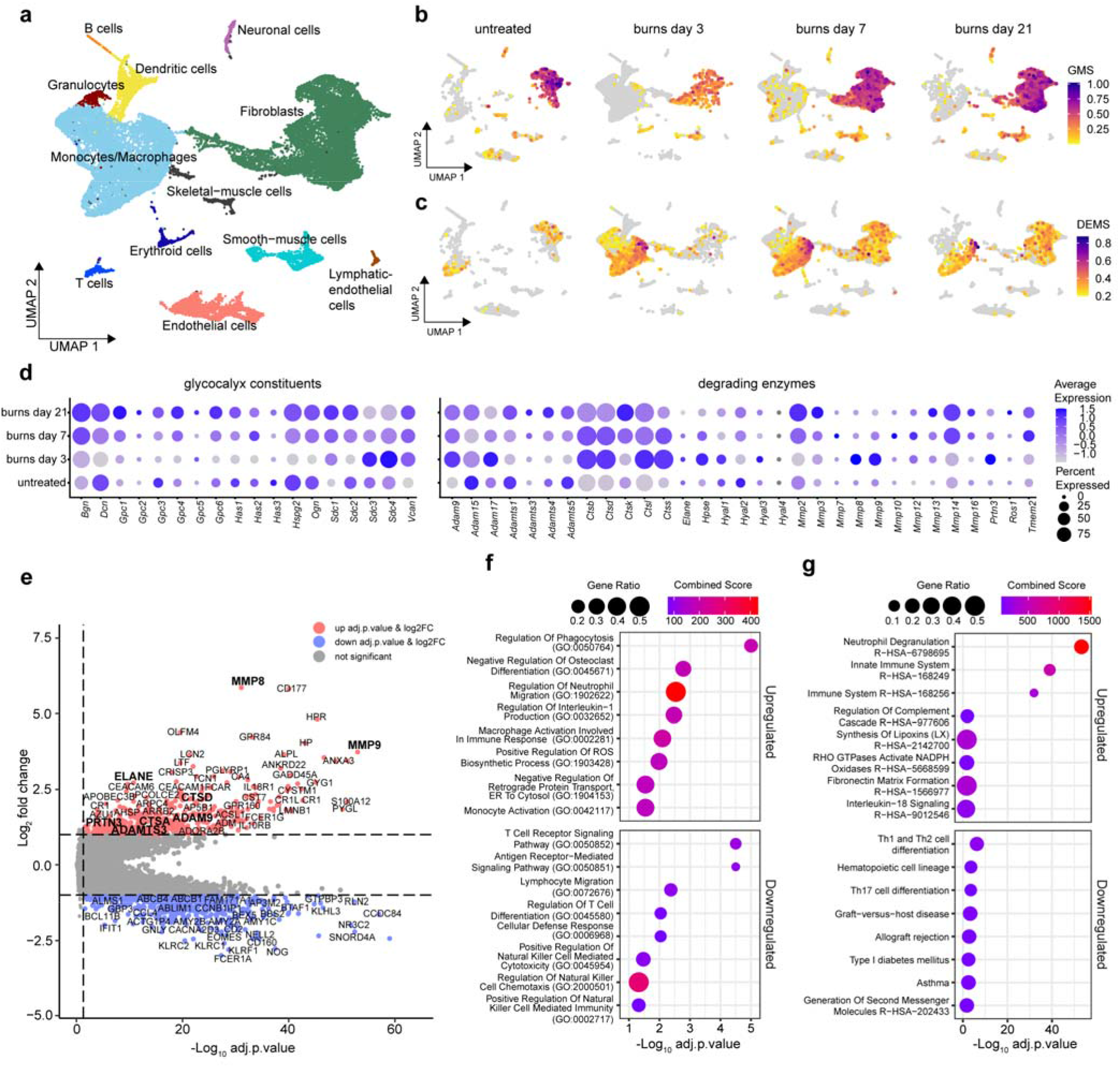
Transcriptomics analysis revealing response of glycocalyx and degrading enzymes post burn trauma. (a) UMAP of comprehensive scRNA-seq analysis identifying 12 unique cell clusters. (b) Feature Plots showing a glycocalyx module score (GMS), split by condition. (c) Feature Plots illustrating a degrading enzyme module score (DEMS) (d) Dot plot of individual glycocalyx constituents and degrading enzymes used for the module scores. Dot size represents the expression percentage for each gene with color intensity reflecting average gene expression levels. (e) Differentially expressed genes (DEGs) between control and burn groups. Significantly (log2FC> 1, adj. p < 0.05) upregulated genes in the burn group show a positive fold change (red), while those downregulated show a negative fold change (blue). (f) The top 8 ‘GO Biological Process 2023’ terms associated with significantly upregulated and downregulated DEGs. (g) The top 8 terms from a combined query of ‘KEGG 2021 Human’ and ‘Reactome 2022’. Results are displayed as dot plots with their respective combined scores and gene ratios.

To extend these murine findings to human systems and enhance their translational relevance, we analyzed transcriptomic data from the “Inflammation and the Host Response to Injury Large-Scale Collaborative Research Program,” which included whole blood samples from 244 severely burned patients with over 20% total body surface area (TBSA) and 35 healthy controls ^13^. The analysis identified several upregulated mRNAs in burn patients involved in neutrophil migration and activation, such as CD177, MPP1, FUT7, RAC2, FPR2, OLFM4, ELANE, MPO, PADI4, and ROMO1. Conversely, genes related to T cell activation and differentiation, including ITK, CD3D, CD3G, ZAP70, and MALT1, were downregulated, indicating an altered adaptive immune response following burn injury (Fig. 1e). Gene set enrichment analysis of significantly regulated genes highlighted enhanced activation of innate immune cells post-burn, focusing on processes such as neutrophil activity, phagocytosis, and macrophage activation. (Fig1f,g). A list of differentially expressed genes between burn patients and controls as well as the genes associated with enriched terms are supplied in supplementary table 1. (Table S1)

While the literature presents conflicting reports on neutrophil migration following burn injury - some studies suggest impairment ^14^, while others indicate enhanced recruitment ^15^ - our analysis revealed an upregulation of genes involved in monocyte, macrophage, and neutrophil activity and migration. The infiltration of neutrophils into the alveoli, a hallmark of ARDS ^6^, directly impacts mortality by increasing lung epithelial and endothelial permeability through mediators such as neutrophil elastase ^16^.

Further analysis revealed that many degrading enzymes, including MMP8, MMP9, ADAM9, HPSE, CTSD, CTSG, ELANE, and PRTN3, were significantly upregulated in burn patients, with MMP8 showing the highest overall log2 fold change of 5.86 (Fig. 1d,e). These findings underscore the intricate link between systemic innate immune activation and glycocalyx disruption, highlighting the heterogeneous responses of glycocalyx components and degrading enzymes. MMP8, in particular, appears to be a conserved and significantly upregulated enzyme following burn injury, potentially contributing to glycocalyx degradation.

To further investigate the systemic effects of burn injury on glycocalyx shedding and MMP8 production, we measured serum levels of syndecan-1 (SDC1), syndecan-4 (SDC4), hyaluronic acid (HA), and MMP8 in burn patients over a 21-day period, comparing them with healthy controls (Fig. 2). For this study, we enrolled 28 burn injury patients and 8 healthy controls (Table S2). Our findings showed that SDC1 levels were consistently elevated from day 1 to day 21 post-burn compared to controls (Fig. 2a). In contrast, HA and SDC4 serum levels exhibited smaller increases that did not reach statistical significance (Fig. 2b,c).

**Figure 2.**
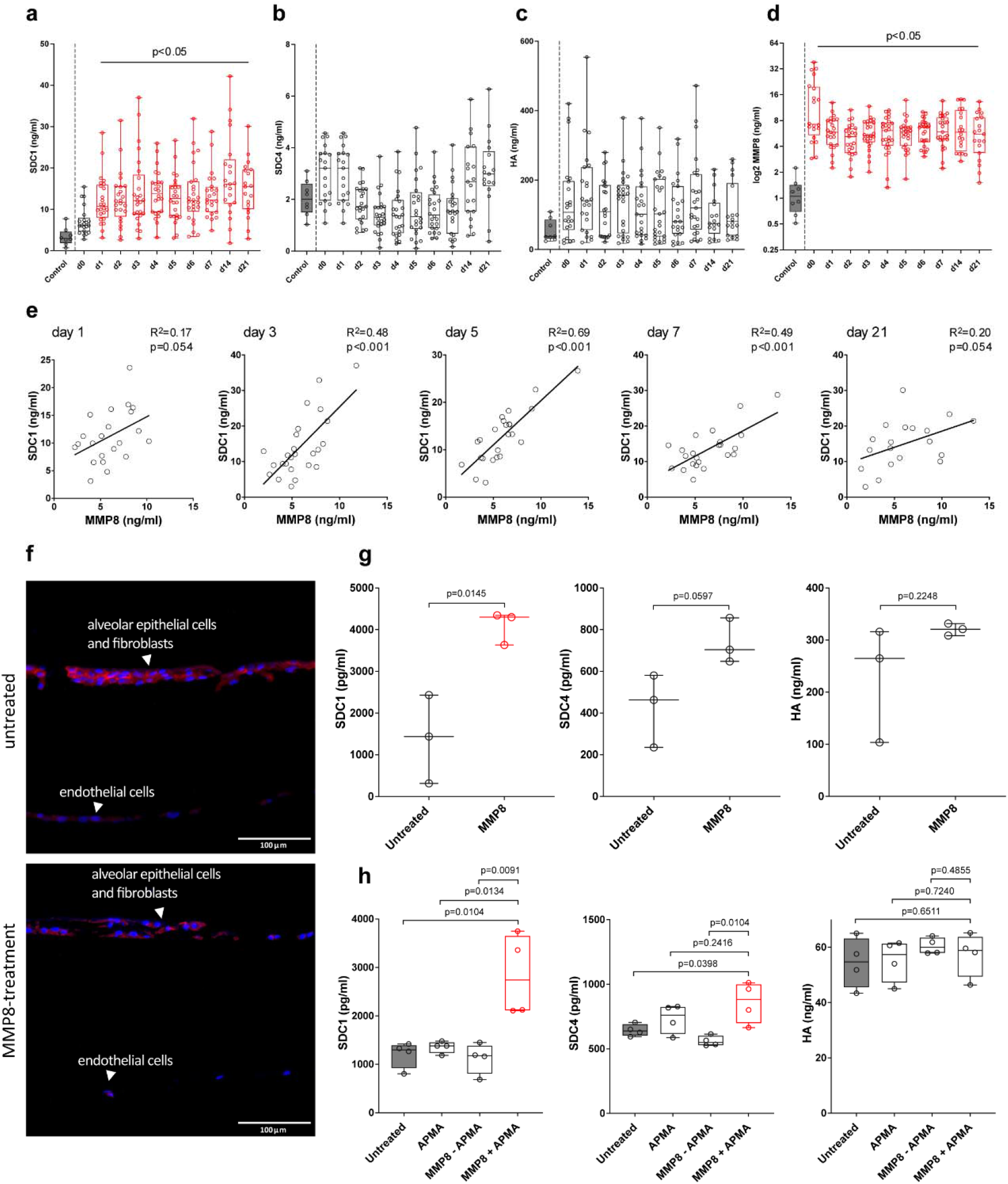
Serum levels of glycocalyx constituents and MMP8 as well as MMP8 effect on glycocalyx shedding in-vitro. Systemic levels of SDC1 (a), SDC4 (b), HA (c) and MMP8 (d) were quantified in healthy controls and burn patients over a 21-day follow-up. Data is presented as minimum to maximum boxplots including all data points, with significant time points marked in red. (e) Pearson correlation analysis between MMP8 and SDC1 serum levels in burn patients over various time points. (f) Immunofluorescence imaging of untreated and rhMMP8-treated EpiAlveolar™ 3D lung models. 20x magnification images display DAPI-stained nuclei (blue) and SDC1 fluorescence (red). (g) SDC1, SDC4, and HA levels in EpiAlveolar™ model supernatants, with significant differences (p < 0.05) marked in red. (h) SDC1, SDC4, and HA supernatant levels in SAECs treated with APMA, rhMMP8 alone, and activated rhMMP8.

Interestingly, while there was no consistent correlation between SDC1 serum levels and the Abbreviated Burn Severity Index (ABSI), Total Body Surface Area (TBSA), or third-degree burns over the 21-day period, inhalation injury was associated with increased odds of elevated SDC1 serum levels in the early post-injury phase (Table S3). This observation is consistent with studies in both porcine and human models, which have shown that inhalation injury increases SDC1 serum levels, a factor linked to endotheliopathy and abnormal fibrinolysis post-burn^17^. Notably, SDC1 levels remained significantly elevated in burn patients compared to healthy controls throughout the 21-day follow-up, suggesting that systemic inflammation and subsequent shedding by immune cell-derived enzymes play roles beyond direct lung damage. Our cohort of burn patients also demonstrated significantly increased MMP8 serum levels over the follow-up period, indicating prolonged activation and degranulation of innate immune cells (Fig. 2d). Notably, MMP8 and SDC1 serum levels showed significant correlation at various time points (Fig.2e) These results suggest that glycocalyx disruption post-burn is a complex process involving both direct lung injury and systemic degradation by various enzymes.

To test whether MMP8 directly cleaves SDC1 in the lung, we treated EpiAlveolar™ human airway 3D tissue models with recombinant human MMP8. Immunofluorescence staining of SDC1 in lung equivalents showed reduced SDC1 presence and morphological changes in the alveolar barrier. Consistent with these observations, MMP8 treatment of 3D lung models led to decreased SDC1 levels in both the epithelium and endothelium. Although the epithelium still showed some SDC1 staining, suggesting rapid re-synthesis, the endothelial integrity was significantly compromised, with lower SDC1 expression, indicating slower regeneration post-treatment (Fig.2f). Quantification of SDC1, SDC4, and HA levels in the supernatants of MMP8-treated 3D models and small airway epithelial cells revealed increased levels of SDC1 and SDC4 following MMP8 treatment (Fig.2g,h).

Building on our observations of MMP8’s involvement in glycocalyx shedding, it is important to highlight that other related MMPs, such as MMP2, MMP3, MMP7, MMP9, and MT1-MMP, are also known to cleave SDC1 and SDC4 ^8^, as they interact with conserved cleavage sites on syndecans. Our findings suggest that MMP8 plays a significant role in syndecan shedding, and to fully understand its specific contribution to pulmonary glycocalyx degradation following burn injuries, further research is needed. In particular, studies that explore MMP8 knockdown could reveal whether other enzymes compensate for its absence, thereby clarifying MMP8’s unique role in this process.

In developing effective treatment strategies for ARDS, an approach that reduces immune cell mobilization and activation while preserving the body’s defense mechanisms is crucial ^6^. Selectively targeting MMP8 could help limit epithelial glycocalyx disruption without compromising host defense, potentially reducing the risk of pulmonary complications in burn patients. For instance, the broad-spectrum MMP inhibitor doxycycline has shown promise in mitigating glycocalyx shedding and improving lung function in models of bleomycin-induced lung injury ^18^. Furthermore, MMP inhibition has demonstrated protective effects against pulmonary damage in endotoxin-induced ARDS in mice ^19^

However, while targeting MMPs appears to be a promising therapeutic strategy, it is essential to recognize that the roles of MMPs in alveolar damage and repair are context-dependent and vary by subtype ^20^. To advance our understanding of post-burn lung injury, future research should focus on the specific roles of individual MMPs, including MMP8, and how they interact with other factors. This deeper insight will be critical for advancing knowledge of pulmonary glycocalyx derangements in burn patients and for guiding the development of more effective therapeutic interventions.

## Data Availability

All scRNA-seq and Affymetrix datasets analyzed in the present study can be publicly accessed via the GEO repository with the identifiers GSE126060 and GSE37069.

https://www.ncbi.nlm.nih.gov/geo/query/acc.cgi?acc=GSE126060

https://www.ncbi.nlm.nih.gov/geo/query/acc.cgi?acc=GSE37069

## Acknowledgements

The authors would like to thank Hans Peter Haselsteiner and the CRISCAR Familienstiftung for their ongoing support of the Medical University/Aposcience AG public private partnership aiming to augment basic and translational clinical research in Austria/Europe. This work was supported by the Austrian Research Promotion Agency (#852748, #862068) and the Vienna Business Agency (#2343727) awarded to HJA. MM was funded by Aposcience AG.

## Author Contributions

Conceptualization: HK, DB, CA, CR, HJA, MM; Data Curation: HK, DB, MS, TH, CSM, MTL, HJA, MM; Formal Analysis: HK, DB MS, HJA, MM; Funding Acquisition: HJA, MM; Investigation: HK, DB, HJA, MM; Supervision: HJA, MM; Writing - Original Draft Preparation HK, DB, MS, TH, CSM, MTL, CA, CR, HJA, MM

## Supplementary material

### Supplementary Figure 1 (Fig.S1)

**Figure S1.**
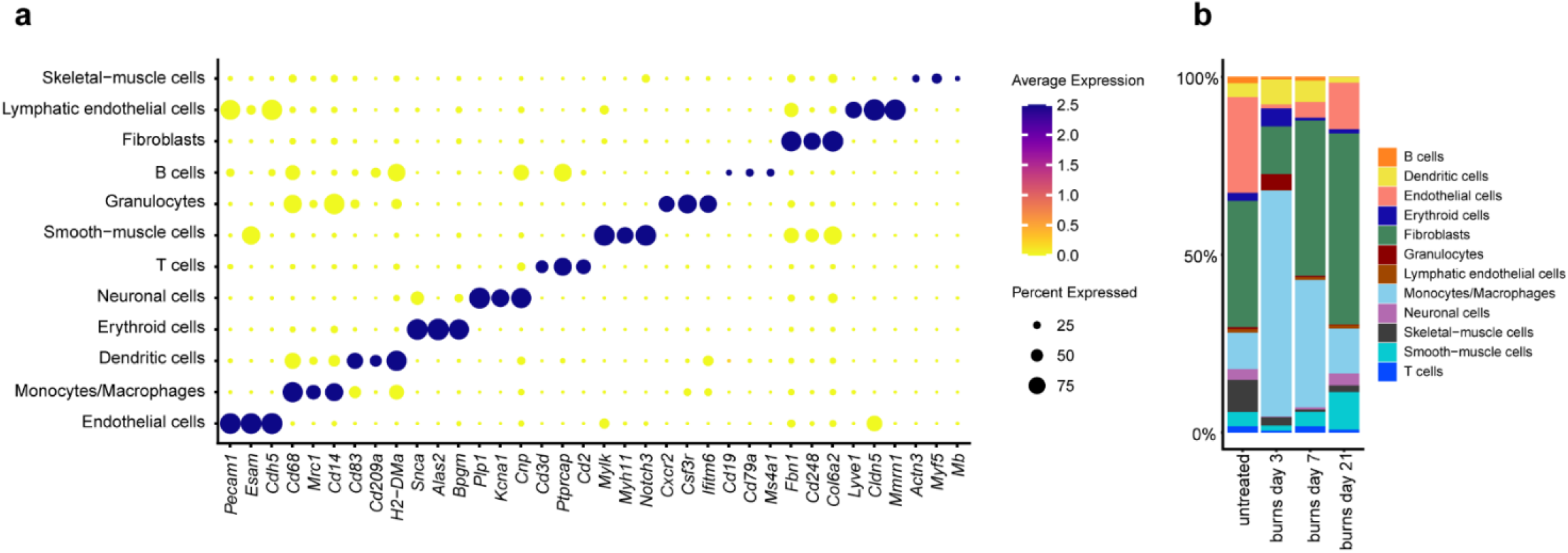
Clustermarkers and cell-type proportions (a) The left panel shows a dot plot visualization of selected cluster-specific markers, with color shading indicating average gene expression and the size of dots representing the percentage of cells expressing each marker. (b) Stacked bar charts on the right panel illustrate the proportional distribution of each cell cluster across different conditions.

### Supplementary table 1 (Table S1)

**Supplementary table 1 (Table S1)** - Differentially expressed genes (DEGs) between control and burn groups and gene set enrichment analysis (GSEA)

⟶ Table S1 is provided as a separate Excel sheet in the Table.S1.xlsx file.

### Supplementary table 2 (Table S2)

**Table S2.** Study population demographics. F:M = female to male ratio, ABSI = Abbreviated Burn Severity Index, APACHE 2 = Acute Physiology and Chronic Health Evaluation II (APACHE 2), LOH = Length of Hospitalization, SAPS II/III = Simplified Acute Physiology Score II/III, TBSA = Total Body Surface Area, LOH = Length of Hospitalization. Indicated are mean (median) ± SD [interquartile range]. *Indicated are n (SD).

| Variable | Burn patients | Controls |
| --- | --- | --- |
| n | 28 | 8 |
| Age (years) | 49.6 (42.5) ± 21.8 [32,25 - 72,75] | 40.5 (36) ± 19.9 [23 - 55,75] |
| F:M ratio (%) | 7: 21 (25:75) | 3:5 (37.5:62.5) |
| ABSI | 7.7 (8) ± 2.8 [5-9] |  |
| APACHE 2 | 18.4 (18) ± 8.3 [11,5 - 26,5] |  |
| LOH (days) | 41.1 (33) ± 34.0 [15 - 67,75] |  |
| SAPS II | 38.3 (35) ± 16.9 [24,75 - 48,5] |  |
| SAPS III | 31.7 (31.5) ± 10.8 [23 - 37,25] |  |
| TBSA (%) | 32.5 (30) ± 20.2 [16,25 - 39,50] |  |
| Deceased (%)* | 3 (10.7) |  |
| Inhalation injury* | 6 (21.4) |  |
| 3rd degree burn (%)* | 19 (67.9) |  |

### Supplementary table 3 (Table S3)

**Table S3.** Correlations between SDC1 and clinical features. The associations between SDC1 serum levels and clinical scores and features in burn patients from admission (d0) to the end of follow-up (d21) are shown. Pearson correlation coefficients were determined for the correlation between two metric variables, as in *r* (ABSI/SDC1) and *r* (TBSA/SDC1). To examine relationships between SDC1 and dichotomous variables (inhalation injury and 3rd-degree burn), simple logistic regression and odds ratio calculations were performed. The table presents the Pearson correlation coefficient (r) or odds ratio (OR), the corresponding 95% confidence interval, and the p-value. Values of statistical significance are highlighted in bold.

| Days after burn | ABSI |  |  | Inhalation Injury |  |  | TBSA |  |  | 3rd degree burn |  |  |
| --- | --- | --- | --- | --- | --- | --- | --- | --- | --- | --- | --- | --- |
|  | Pearson r | 95% CI | p-value | OR | 95% CI | p-value | Pearson r | 95% CI | p-value | OR | 95% CI | p-value |
| **0** | 0.37 | -0.07 - 0.69 | 0.097 | **1.91** | **1.25 - 4.16** | **0.001** | 0.17 | -0.28 - 0.56 | 0.462 | 1.23 | 0.90 - 1.95 | 0.212 |
| **1** | 0.27 | -0.15 - 0.61 | 0.202 | **1.47** | **1.12 - 2.68** | **0.002** | 0.04 | -0.37 - 0.43 | 0.862 | 1.19 | 0.98 - 1.56 | 0.087 |
| **2** | **0.62** | **0.29 - 0.82** | **0.001** | **1.58** | **1.15 - 2.88** | **<0.001** | **0.54** | **0.18 - 0.78** | **0.006** | 1.15 | 0.98 - 1.43 | 0.106 |
| **3** | **0.50** | **0.13 - 0.75** | **0.011** | **1.39** | **1.13 - 2.00** | **<0.001** | 0.29 | -0.12 - 0.61 | 0.161 | 1.12 | 0.99 - 1.35 | 0.091 |
| **4** | **0.46** | **0.07 - 0.73** | **0.023** | **1.42** | **1.11 - 2.17** | **0.002** | 0.30 | -0.12 - 0.63 | 0.158 | 1.20 | 1.00 - 1.55 | 0.056 |
| **5** | 0.29 | -0.12 - 0.62 | 0.164 | **1.31** | **1.06 - 1.84** | **0.009** | 0.28 | -0.13 - 0.60 | 0.182 | 1.14 | 0.96 - 1.41 | 0.138 |
| **6** | -0.04 | -0.42 - 0.36 | 0.860 | 1.08 | 0.94 - 1.24 | 0.266 | 0.00 | -0.39 - 0.39 | 0.991 | 1.03 | 0.91 - 1.20 | 0.636 |
| **7** | -0.05 | -0.45 - 0.37 | 0.832 | 1.11 | 0.93 - 1.35 | 0.250 | 0.05 | -0.37 - 0.45 | 0.817 | 1.04 | 0.88 - 1.29 | 0.662 |
| **14** | 0.04 | -0.40 - 0.46 | 0.874 | **1.14** | **1.02 - 1.33** | **0.022** | -0.09 | -0.50 - 0.35 | 0.695 | 0.99 | 0.89 - 1.12 | 0.844 |
| **21** | 0.14 | -0.33 - 0.56 | 0.564 | 1.16 | 0.98 - 1.48 | 0.090 | 0.06 | -0.40 - 0.50 | 0.792 | 0.95 | 0.79 - 1.13 | 0.588 |

