## Supplementary material for "The impact of burn trauma on glycocalyx derangement": Materials and Methods

Ethical Statement

This study was approved by the institutional review board of the Medical University of Vienna (vote 1997/2023) and was conducted adhering to the principles outlined in the Declaration of Helsinki, as well as in compliance with the “Good Scientific Practice” guidelines by the Medical University of Vienna

Patient Enrollment and Sample Collection

For this study’s investigation, serum samples that have been previously assessed for sST2 and IL-33 levels, as well as markers for neutrophil activation, were examined.^1,2^ A total of 28 patients and 8 control subjects were enrolled in the study. Patients admitted to the Intensive Care Unit 13i1 and the Department of Plastic and Reconstructive Surgery at the Medical University of Vienna within 24 hours of burn trauma, aged over 18 years, and with injuries covering at least 10% of the Total Body Surface Area (TBSA) met the inclusion criteria. The accurate determination of TBSA was achieved by marking the affected areas on a blank human body diagram and subsequent quantification during the initial clinical examination. Individuals presenting with either chronic infectious diseases or an autoimmune disorder were deemed ineligible for this study. The clinical treatment of the patients remained unaltered throughout their participation in the study. During routine daily blood withdrawals in the first week after hospital admission, an additional 27ml was collected for research purposes, followed by weekly sampling for the remainder of the follow-up. Mortality was defined as any instance of all-cause mortality occurring within the period of hospitalization. Patients with third-degree burns were categorized as 'third-degree positive.' This classification was applied irrespective of the extent of the burn-affected region. Eight healthy volunteers were enrolled for comparative analysis to serve as the control group. The blood samples from both the control and patient subjects were processed in accordance with a consistent protocol for both groups. Following a thirty-minute incubation time at room temperature, the whole blood was centrifuged at 2850 g for 17 minutes for serum separation. To ensure stability for subsequent analysis, the obtained samples were stored at temperatures below -70 °C to -80 °C.

Quantification of Serum Analytes via Enzyme-Linked Immunosorbent Assay

The assessment and quantification of serum Syndecan 1 (SDC1), Syndecan 4 (SDC4), Hyaluronan (HA), and Matrix Metalloproteinase-8 (MMP-8) was conducted via enzyme-linked immunosorbent assay (ELISA). Following commercially available ELISA kits were used, adhering to the manufacturers’ instruction: Human Syndecan-1 DuoSet ELISA kit (R&D Systems®, cat#: DY2780), Human Syndecan-4 DuoSet ELISA kit (R&D Systems®, cat#: DY2918), Hyaluronan DuoSet ELISA kit (R&D Systems®, cat# DY3614-05) and Human Total MMP-8 DuoSet ELISA kit (R&D Systems®, cat#: DY908). Appropriate dilution of the burn serum samples for the ELISA assay was performed as follows: Syndecan 1 (1:10), Syndecan 4 (1:10), Hyaluronan (1:5), MMP-8 (1:20). Supernatants from in-vitro were used undiluted, except for Hyaluronan, which was diluted 1:5. The optical density readings were obtained by using a Tecan F50 infinite microplate reader (Tecan Group, Männedorf, Switzerland) and processed via Magellan software version (Tecan Group, Männedorf, Switzerland). Analyte concentrations were deduced from external standard curves.

Statistical Analysis

For statistical analysis and visualization of the data, IBM SPSS Statistics version 29.0.0 (IBM, Armonk, NY, USA) and GraphPad Prism version 10.0.3 (GraphPad Software, San Diego, CA, USA) were used. The Enzyme-Linked Immunosorbent Assay (ELISA) data was preprocessed with the application of the Robust regression and Outlier removal (ROUT) method (Q=1%). After the detection and removal of outliers, a Shapiro-Wilk test was performed to evaluate the normality of data distribution. Data not adhering to Gaussian distribution prerequisites, was further analysed using Kruskal-Wallis with Dunn’s multiple comparison tests. In the case of the in vitro experiments, comparisons between two groups with normally distributed data were performed using unpaired t-tests. For correlation analysis between two metric variables, Pearson correlation coefficients were calculated. Additionally, simple logistic regression analyses and odds ratio calculations were conducted to examine associations between nominal and metric variables. The ELISA results are presented as minimum-to-maximum boxplots showing all data, with significant time points (adjusted p-value <0.05) distinctly marked in red.

Single-cell RNA sequencing (scRNA-seq) Analysis

Publicly available single-cell data from the NCBI Gene Expression Omnibus (GEO) database (GSE126060) were retrieved. The data were derived from a study involving a murine burn-tenotomy model.^3^ Briefly, animals received a partial-thickness burn injury covering 30% of their total body surface area (TBSA) on their dorsum, together with an Achilles tendon transection on the hind limb. Tissue samples were collected from the hind at day 0 (no burn/tenotomy) and on days 3, 7, and 21 post-injury. Samples were then subjected to single-cell RNA sequencing analysis. Computational analysis of single-cell RNA sequencing (scRNA-seq) was conducted using R (version 4.3.1, The R Foundation, Vienna, Austria) and RStudio (version 2023.6.2.0 + 561). The Seurat package (version 4.9.9.9059) was used for analysis, adhering to the developer’s Seurat v5 workflow for exploratory data analysis and quality control.^4^ In the data preprocessing stage, Seurat objects were merged into a single object and subjected to quality control. Cells with less than 400 or more than 40,000 unique molecular identifiers (UMIs), expressing fewer than 200 or more than 6,000 genes, and cells with over 10% mitochondrial reads were excluded from downstream analysis. In addition, mitochondrial genes and genes with less than three UMI counts per feature were removed from the UMI count matrices. For data integration, the merged dataset was divided into 'count' and 'data' layers, followed by normalization and the identification of variable features, as outlined in the Seurat v5 integrative analysis guidelines. Prior to Principal Component Analysis (PCA) for dimensionality reduction, features were scaled and centered. Data integration was performed using the “IntegrateLayers” function with an anchor-based RPCA approach, as per Seurat v5 vignette. The resulting single, integrated, batch-corrected expression matrix formed the basis for all further analyses. Clustering was done according to the standard Seurat workflow, which included UMAP (Uniform Manifold Approximation and Projection) and Louvain clustering through the “RunUMAP,” “FindNeighbors,” and “FindClusters” functions. The first 25 principal components (PCs) with a clustering resolution set at 0.3 were utilized. The identification of differentially expressed genes (DEGs) was conducted using the MAST statistical framework ^5^, incorporated in Seurat's “FindMarkers” and “FindAllMarkers” functions, focusing on genes expressed in a minimum of 25% of cells in one group. The "AddModuleScore" function in the Seurat environment was utilized to calculate module scores for gene sets. For the glycocalyx module score (GMS),expression levels of *Bgn, Dcn, Gpc1-6, Has1-3, Hspg2, Ogn, Sdc1-4* and *Vcan* genes were incorporated. The degrading enzyme module score (DEMS) included expression levels of *Adam9, Adam15, Adam17, Adamts1, Adamts3, Adamts4, Adamts5, Ctsb, Ctsd, Ctsk, Ctsl, Ctss, Elane, Hpse, Hyal1, Hyal2, Hyal3, Hyal4, Mmp2, Mmp3, Mmp7, Mmp8, Mmp9, Mmp10, Mmp12, Mmp13, Mmp14, Mmp16, Prtn3, Ros1* and *Tmem2* genes.

DNA Microarray and Gene Set Enrichment Analysis

Human Affymetrix U133 Plus 2.0 GeneChip™ data (GSE37069) were retrieved from the NCBI's GEO database, employing GEOquery version 2.66.0 ^6^ for the download. Differential gene expression (DEG) analysis was performed via the limma package version 3.54.0 ^7^ in R (version 4.3.1) using RStudio (version 2023.6.2.0 + 561). For our analysis, DEGs between the control and burn groups with a log2fold change >1 and an adjusted p-value <0.05, following the Benjamini & Hochberg correction method, ^8^ were considered significant. These DEGs were functionally annotated using the Enrichr ^9^ package, accessing “GO Biological Process 2023”, “KEGG 2021_Human,” and “Reactome 2022” databases. Results from Enrichr were refined by filtering terms with a Benjamini-Hochberg adjusted p-value below 0.05. These were then ranked based on the "combined score,", which incorporates Fisher’s exact test p-values and rank deviations. The enriched terms are illustrated in dot plots, depicting combined score and gene ratios of genes.

Bioinformatics Data Visualization

Bioinformatics analysis results were visualized using the following R packages: Seurat v. 4.9.9.9059 ^4^, ggplot2 v.3.4.2 ^10^, EnhancedVolcano v.1.18.0 ^11^, and scCustomize v.1.1.1 ^12^.

Cell Culture and MMP-8 In-Vitro Assay

EpiAlveolar^TM^ 3D Small Airway Human MicroTissues, incorporating endothelial cells as well as primary human fibroblasts and alveolar epithelial cells cultivated at an air-liquid interface were purchased from MatTek (MatTek Corporation, Massachusetts, USA, cat#: ALV-100-MM)

3D models were maintained using 5 mL of Alveolar assay/maintenance medium (MatTek Corporation, Massachusetts, USA, cat#: ALV-100-MM) in the basal compartment and 75 µl on the apical surface, in accordance with the manufacturer’s instructions. The inserts were placed in 12-well plates for subsequent treatments with activated Recombinant Human MMP-8 Protein (R&D Systems®, cat#: 908-MP-010). Clonetics™ Human Small Airway Epithelial Cells (SAECs), (Lonza Group AG, Basel, Switzerland, cat#: CC-2547) were cultured in SABM™ Basal Medium (Lonza Group AG, Basel, Switzerland, cat#: CC-3119) supplemented with SAGM™ SingleQuots™ Kit Supplements & Growth Factors (Lonza Group AG, Basel, Switzerland, cat#: CC-4124). Cells were treaed with Recombinant Human MMP-8 Protein, CF (R&D Systems®, cat#: 908-MP-010) at confluency. Recombinant human MMP-8 (rhMMP-8) was activated at a concentration of 100 µg/mL using 1 mM p-Aminophenylmercuric Acetate (APMA), (Sigma-Aldrich®, cat#: 164610-700MG) in a buffer containing 50 mM Tris, 10 mM CaCl_2_, and 150 mM NaCl (pH 7.5). The activation reaction was incubated at 37 °C for 1 hour. Following activation, the SAECs were treated with the activated rhMMP-8 at 1.0 ng/µl concentration in SABM™ Growth medium. As part of the experimental design, SAECs were also treated with inactive rhMMP-8 (prepared without adding APMA), APMA alone in SABM™ Growth medium, or with the growth medium alone as a control. EpiAlveolar^TM^ 3D Small Airway Human MicroTissues were treated with activated rhMMP-8 in alveolar assay medium supplied by the manufacturer or for control with the alveolar assay medium alone.

Immunofluorescence Staining and Microscopy

Sections embedded in paraffin were deparaffinized using a heater fan until the paraffin melted, ensuring the slides did not dry out at any point. The slides were then treated with xylene and sequentially rehydrated in ethanol solutions (96%, 80%, 30%). After rehydration, the sections were rinsed twice in deionized water (dH2O) followed by antigen retrieval using a 10mM sodium citrate buffer (pH 6). Sodium citrate buffer was prepared by diluting 25mL of Target Retrieval Solution 10x (Dako, cat#: S2369) in 225mL dH2O. Subsequently, slides were placed in a jar containing buffer and heated in a pressure cooker for 20 minutes, followed by cooling to room temperature. Prior to staining, the sections were washed in Phosphate-Buffered Saline (PBS). The primary Anti-Syndecan-1 antibody (Abcam, Cambridge, UK, cat#: ab128936)was diluted 1:500 in 2% PBS/BSA, applied to each section, and incubated overnight at 4°C. Post-incubation, the sections were washed with PBS and treated with a red fluorescent secondary antibody, Goat anti-Rabbit IgG (H+L) Alexa Fluor™ 546 Invitrogen, ThermoFisher Scientific, cat#: A-11035), diluted 1:500 in 2% PBS/BSA, supplemented with 10% goat serum and DAPI (1:1000) (ThermoFisher Scientific, cat#: 62248). After three washes in PBS and dH2O, the slides were mounted using Aqua Polymount medium (Polysciences, Warrington, PA, USA, cat#: 18606) and stored in the refrigerator. Fluorescence microscopy was performed on an OLYMPUS BX63 microscope, with images captured and analyzed using Olympus cellSens software (Olympus, Shinjuku, Tokyo, Japan).
